# Peak Agricultural Pesticide Spray Seasons Are Associated with worse Negative Affect and Behavioral Dysregulation in Adolescents and Young Adults: A 2-Year Ecological Momentary Assessment Study in ESPINA

**DOI:** 10.64898/2026.10.14.26365000

**Authors:** Briana N.C. Chronister, Emma M. Parrish, Dolores Lopez-Paredes, Jose Suarez-Torres, Avery Quynh, Danilo Martinez, Asa Bradman, Raeanne C. Moore, Jose R. Suarez-Lopez

**Affiliations:** Herbert Wertheim School of Public Health and Human Longevity Science, University of California San Diego, La Jolla, CA, 92093, USA; Department of Veterans Affairs Medical Center, San Diego, CA, USA; Department of Psychiatry, University of California San Diego, La Jolla, CA, 92093, USA; Fundación Cimas del Ecuador, Quito, Ecuador; Department of Public Health and Environmental Systems Graduate Group, University of California, Merced, CA, USA; Department of Pediatrics, University of California San Diego, San Diego, 92123

**Keywords:** Pesticides, Mood, Positive Affect, Agriculture, Youth

## Abstract

**Introduction:** Pesticide exposure has been associated with poorer mental health, yet little is known about whether seasonal fluctuations in agricultural pesticide use are associated with cyclical changes in mood. We evaluated associations between 10 peak and off-peak pesticide spray seasons and ecological momentary assessment (EMA)-derived mood in a floricultural region in Ecuador.

**Methods:** 496 adolescent and young adult participants completed repeated EMAs over 2.5 years, including one single-assessment administration and nine 10-day bursts (N_assessments_=14,332). Mood outcomes included negative affect (sadness, anxiety, stress, anger; overall mean [SD]=2.79 [1.32]), positive affect (happiness, motivation; mean=4.21 [1.60]), behavioral dysregulation (aggression and impulsivity; mean=1.86 [1.29]), and fatigue and concentration (tiredness and difficulty concentrating; mean=3.25 [1.34]). Generalized linear mixed models estimated pooled, between-person and within-person associations between peak pesticide spray seasons and mood, adjusting for demographic and longitudinal factors. Effect modification by sex, floricultural occupation, and cohabitation with a floricultural worker was assessed.

**Results:** Peak spray seasons were associated with higher negative affect (β=0.11 [0.06, 0.16]) and behavioral dysregulation (β=0.10 [0.05, 0.15]), fatigue and difficulty concentrating (β=0.06 [0.001, 0.12]), but not positive affect (β=0.01 [−0.05, 0.07]), Associations were driven by within-person changes. Sex modified associations with anger, which were stronger in males than females. Floricultural occupation modified associations, with greater behavioral dysregulation and lower positive affect among floricultural workers than non-workers.

**Discussion:** Peak pesticide spray seasons were associated with worsening negative affect, behavioral dysregulation, and fatigue and concentration. EMA may provide a sensitive approach for detecting short-term neurobehavioral changes associated with seasonal pesticide exposure.

## Introduction

Mental health conditions can reduce life expectancy by 10 years, and contribute up to 8 million deaths worldwide annually^1^. Reported median age of onset is approximately 31 years (Interquartile Range=21-46 years) for mood disorders, 30 years (21-44 years) for depressive disorders, and 32 years (20-42 years) for generalized anxiety disorder ^2^. Adolescents and adults with anxiety and depression are at higher risk of adverse psychosocial outcomes, including increased suicidal ideation and behavior ^3,4^, social functioning problems ^5^, and poor health ^5^. The tripartite model of anxiety and depression provides a useful framework for understanding the connection of these conditions and mood, demonstrating that they share a common dimension of elevated negative affect, that reduced positive affect is more characteristic of depression and that physiological hyperarousal is more characteristic of anxiety ^6^. With this context, the transition period from adolescence to young adulthood is a crucial life stage to identify contributing factors to the incidence of mental health conditions to inform prevention strategies, as early interventions have been found to improve health outcomes ^7,8^.

Pesticide exposure has been consistently linked to adverse mental health outcomes, including elevated symptoms of depression and anxiety, particularly among occupational and para-occupational populations in agricultural settings. Among occupational workers, farmworkers who experienced pesticide poisonings had 194% higher odds of depression ^9^. A systematic review of 27 articles found 78% of studies reported associations between pesticide exposure and incident depression, with the strongest evidence involving organophosphate insecticides, herbicides, and pyrethroids ^10^. Similar trends have been observed in para-occupational populations (household members of workers with occupational pesticide contact) have been found. For example, female spouses of licensed pesticide applicators had greater odds of experiencing depression^11^. Children who lived with floricultural workers in Ecuador had lower acetylcholinesterase (AChE) activity, indicating greater exposure to organophosphate and carbamate (cholinesterase inhibiting) pesticides within a 82 day window ^12^. In this population, lower AChE activity was associated with higher depression symptoms, with stronger associations observed in girls compared to boys ^13,14^. Similar associations in other populations report that higher exposure was associated with higher hospitalizations rates due to mood disorders ^15^. Individuals living and working near intensive agricultural production may experience chronic, low-level exposure to complex mixtures of commonly used pesticides, such as organophosphates, pyrethroids, neonicotinoids, and herbicides ^16,17^ through take-home pathways ^18–20^, household contamination ^17^, environmental drift ^21^, or water contamination^22^. These associations are biologically plausible because pesticides may influence neurobehavioral functioning through cholinergic dysregulation ^13,23^, oxidative stress ^24,25^, and endocrine disruption ^26^; however, the temporal dynamics of these associations remain poorly understood.

In floricultural regions such as Pedro Moncayo County, Ecuador, pesticide application follows distinct seasonal patterns tied to crop cycles and market demands. Pedro Moncayo exports roses worldwide and houses one of the highest concentrations of flower plantations in the Americas. The region produces up to 25% of the total rose exports from Ecuador, which are shipped to the United States, Russia, and Europe ^27^. Roses are perennial flowers ^28^ whose major harvest periods in Ecuador align with popular northern hemisphere holidays such as Valentine’s day in February, Mother’s Day, which is celebrated in May in ∼63% of the world ^29^, and Christmas in December. As pesticide use intensifies to meet export production demands, rose harvest periods coincide with peak pesticide spray periods. Prior studies have demonstrated that lower AChE activity has been observed during peak pesticide spray seasons in occupational populations ^30–32^, and there is variable exposure by season ^33^. In children living in Pedro Moncayo examined as part of the Study of Secondary Exposures to Pesticides across Childhood, Adolescence and Adulthood (ESPINA), we previously observed positive linear associations between time after the Mother’s Day Flower Harvest and AChE, which were strongest in those who lived within 233m of a flower plantation ^34^. These findings suggest peak pesticide spray seasons put local agricultural communities, occupational workers, and para-occupational populations at increased risk of pesticide exposure. Importantly, in this population we also observed that participants examined sooner after the Mother’s Day Harvest had lower neurocognitive performance in several domains compared to those examined later ^35,36^, which supports the hypothesis that pesticide spray seasons may also result in transient brain health alterations.

Despite variability in pesticide exposure across spray periods, most epidemiologic studies examining pesticide exposure and mental health rely on single or infrequent assessments of mood, typically collected in clinical or laboratory settings. Such approaches fail to capture the dynamic and context-dependent nature of mood; potentially obscuring short-term fluctuations associated with changing environmental exposures. Ecological momentary assessment (EMA) offers a powerful alternative by enabling real-time, repeated measurement of mood in participant’s natural environments ^37,38^ and have been found to outperform paper-and-pencil measures in capturing mood changes ^38^. EMA reduces recall bias and enhances ecological validity, providing a more sensitive approach to detecting transient changes in affect that may correspond with exposure variability. National efforts to improve cellular infrastructure and digital literacy in Ecuador has led to increased mobile phone use ^39^. In 2017, of the national 16-24 year-old population, 69.3% had an active smartphone and 90.1% were digitally literate ^40^.

Since 2020, estimated smartphone ownership among Ecuadorian young adults have ranged from 85% ^41^ to 98% ^42^. Leveraging EMA in this context allows for the characterization of between-person and within-person changes in mood across time and environmental conditions, including periods of intensified pesticide use, providing unique opportunity to examine how fluctuations in environmental exposures translate into real world mood outcomes.

In this study, we evaluated associations between peak pesticide spray seasons and mood assessed using EMA across ten repeated single and burst administrations over a two-year period in a young adult rural population living in Pedro Moncayo, Ecuador. By capturing high-resolution, real-time mood data, this study seeks to advance understanding of the short-term and cumulative mental health impacts of peak pesticide use periods in agricultural populations.

## Methods

### Setting

Pedro Moncayo, Ecuador, nicknamed the Capital of the Rose (population ∼40,000^43^), is known for agricultural production ^44^.This region has an ideal climate for agriculture due to its minimal seasonal variation in climactic factors such as temperature, precipitation levels, day length, and fertile soil, permitting year-round planting of crops ^45^. In 2022, floricultural greenhouses comprised ∼5% of Pedro Moncayo’s total surface area. Production of floriculture occurs year round ^29,46^.

### Study Design

The Cyclical Alterations in Neurocognitive Performance and Mood Over Two Years in Relation to Pesticide Spray Seasons Among Adolescent and Young Adult Participants of the ESPINA study (ESPINA-EMCT) is an ancillary study launched in tandem with the 14^th^ follow-up year (FUY) exam of ESPINA study participants in July 2022 (FUY-14a). Enrollment procedures for the original ESPINA cohort have been previously published ^13,18,47^. ESPINA is a prospective cohort study that has conducted 13 follow-up examinations over 16 years since its inception in 2008, when participants were 4-7 years old. Cumulatively since 2008, ESPINA has examined 751 participants from childhood through young adulthood to assess the health impacts of pesticide exposure across the life course ^18,48,49^. Participants have been recruited from the Survey of Access and Demand of Health Services (SADHS), the System of Local and Community Information (SILC), community announcements, and word-of-mouth.

ESPINA-EMCT spans 10 completed assessments, which occurred once every 2-4 months from July 2022 (FUY-14a) to September 2024 (FUY-16d) (Figure 1). For the FUY-14a assessment cohort (n=505), 386 participants had completed the ESPINA FUY-8b assessment, and 119 participants were newly recruited using the SILC, community announcements, and word-of-mouth like prior assessments. The first assessment (FUY-14a) consisted of a single in-person administration of the study measures at local schools. Assessments 2-5 consisted of remote bursts of daily EMA surveys over 10 days. To reduce participant fatigue and improve compliance, the number of EMA administrations per burst was reduced for administrations 6-10, which consisted of remote bursts administered every other day over 10 days. Participants completed the burst 1 assessment in local schools using study-provided smartphones (Samsung Galaxy A03 Core) connected to a private Wi-Fi network. Subsequent bursts were completed in participants’ real-world settings, such as at home or in indoor/outdoor environments, using personal smartphones, including Android and Apple devices.

**Figure 1.**
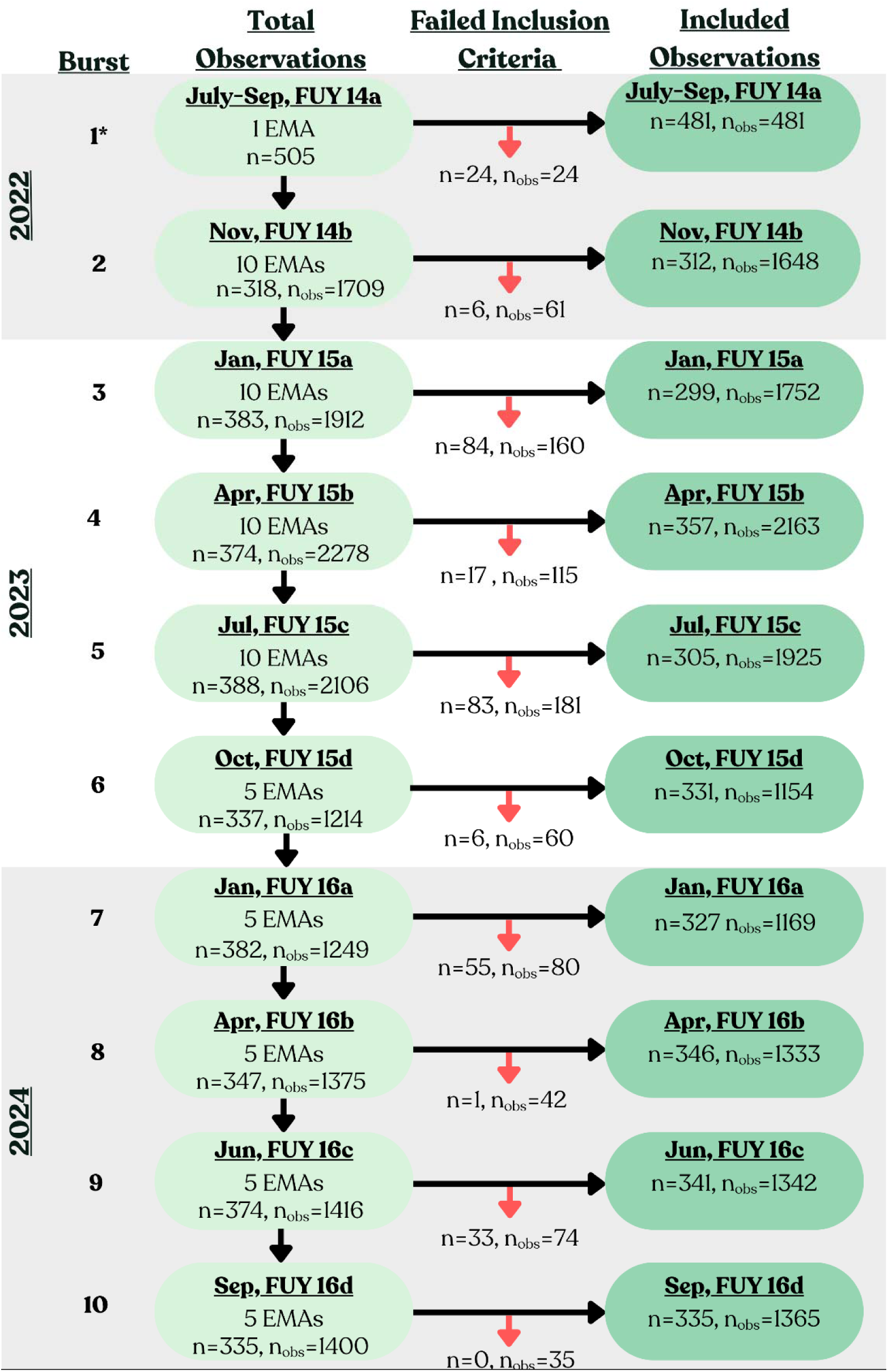
Flow Diagram of Participant Inclusion and Ecological Momentary Assessment Observations Across Follow-Up Bursts for ESPINA-EMCT (July 2022-September 2024). Inclusion criteria included completion of the ecological momentary assessment (EMA), had non-missing covariates, and met alcohol and cannabis inclusion criteria. *Burst 1 included a single administration of the EMA.

This study was approved by the institutional review boards at the University of California San Diego (La Jolla, United States) and UTE University (Quito, Ecuador) and was registered in the Ministry of Public Health of Ecuador.

### Ecological Momentary Assessments

Participants completed a cognitive battery and a brief EMA survey in Spanish using NeuroUX (www.getneuroux.com) weblinks for burst 1 and the native app for Android and iPhone for bursts 2 through 10, which captures data offline. NeuroUX is a regulatory-compliant platform that delivers mobile EMA surveys, and has been used in diverse populations ^50,51^. Each survey took on average 7 minutes to complete. Data collected through the NeuroUX app were temporarily stored locally on the device and automatically uploaded to the NeuroUX platform once an internet connection became available, allowing study staff to access participant data in near real-time. The NeuroUX app was accessible through both the Apple App Store and Google Play Store. To facilitate compliance, the study team sent WhatsApp text message reminders of the upcoming bursts to all participants before the initiation of each burst.

Participants could complete a maximum of one EMA survey during Burst 1, ten EMA surveys during Bursts 2-5, and five EMA surveys for Bursts 6-10 (Figure 1; see *Study Design*). For the first burst, all participants completed the survey once at the study testing sites at local schools primarily on the weekends from 8am to 3pm. For bursts 2-10, the surveys were active from 4am to 11:59 pm on their smartphone apps providing participants a flexible window to complete the assessment at a convenient time. In addition to the WhatsApp text reminders, the NeuroUX app sent up to 3 notifications per day as reminders for participants to complete their daily session.

### Capture and Imputation of Participant Demographics

Participant education, sex, race, and monthly household income were reported once during burst 1. Floricultural occupation and cohabitation with a floricultural worker were assessed at multiple bursts. These variables were reported directly at bursts 1, 7, and 9. At burst 7, participants retrospectively reported their occupation and cohabitation status for bursts 3-7; at burst 9, they retrospectively reported these variables for bursts 7–9.

To maximize observations, we used a rule-based single imputation strategy that leveraged repeated measures from ESPINA and ESPINA-EMCT. After characteristics were properly attributed to each burst, missing data for monthly household income, participant education, occupation, and cohabitation status were imputed using a forward, stepwise approach. First, burst 1 values were imputed using the proceeding ESPINA assessment, FUY-8b. Then, observations were sequentially imputed using data reported from the immediately preceding assessments (e.g., FUY-14b imputed using FUY-14a values).

### Mood Measurements

Each EMA survey included questions regarding current mood on a Likert scale from 1 (not at all) to 7 (extremely). Four questions measured current negative affect: I feel 1) sad or depressed, 2) anxious or nervous 3) stressed, 4) angry or upset. Two questions measured fatigue and concentration: 1) tired, and 2) I’m having trouble concentrating. Two questions measured current positive affect: I feel 1) happy and 2) motivated. Likewise, two questions measured behavioral dysregulation: I feel 1) aggressive and 2) impulsive. Scores for the four mood domains averaged their component questions, thus greater scores indicated worse negative affect, behavioral dysregulation, and fatigue and concentration, but better positive affect. These measures are face-valid questions that have high correlations with the NIH Reported Outcomes Measurement Information System (PROMIS) depression (Cronbach α=0.90) and anxiety (Cronbach α =0.93) short forms ^38^, and have demonstrated sensitivity to changed clinical symptoms in adults ^38^. Each EMA survey also asked about recent alcohol and cannabis use (“How many alcoholic drinks have you had today?” “Have you used marijuana/THC/cannabis today?” Answers of 1 or more prompted follow-up quantity questions).

### Statistical Analysis

Observations were excluded if participants reported any alcohol or cannabis use within the 2 hours prior to the EMA assessment, or four or more alcoholic drinks at any time that day. Descriptive statistics were calculated, and differences between high and low pesticide spray seasons were assessed using independent t-tests for continuous variables, or chi-squared analyses for categorical variables. We used scatterplots to visualize mean mood scores across the 10 bursts; longitudinal associations between the bursts and averaged mood scores by burst were estimated using generalized linear mixed models (GLMM). For these models, the fixed effects included longitudinal burst number (bursts 1–10), and a priori–selected confounders: age, sex, monthly household income (<$500 USD, $500-<$1000 USD, and $1000+), and participant education (years). Burst number was adjusted as a fixed effect to account for systematic longitudinal time trends observed across the duration of the study (Figure 2), plausibly resulting from participant fatigue, habituation, or contextual changes between adolescence to young adulthood (e.g. higher stress due to joining the work force, higher education, etc.). To account for the hierarchical structure of the data, random effects included random intercepts for participant.

**Figure 2.**
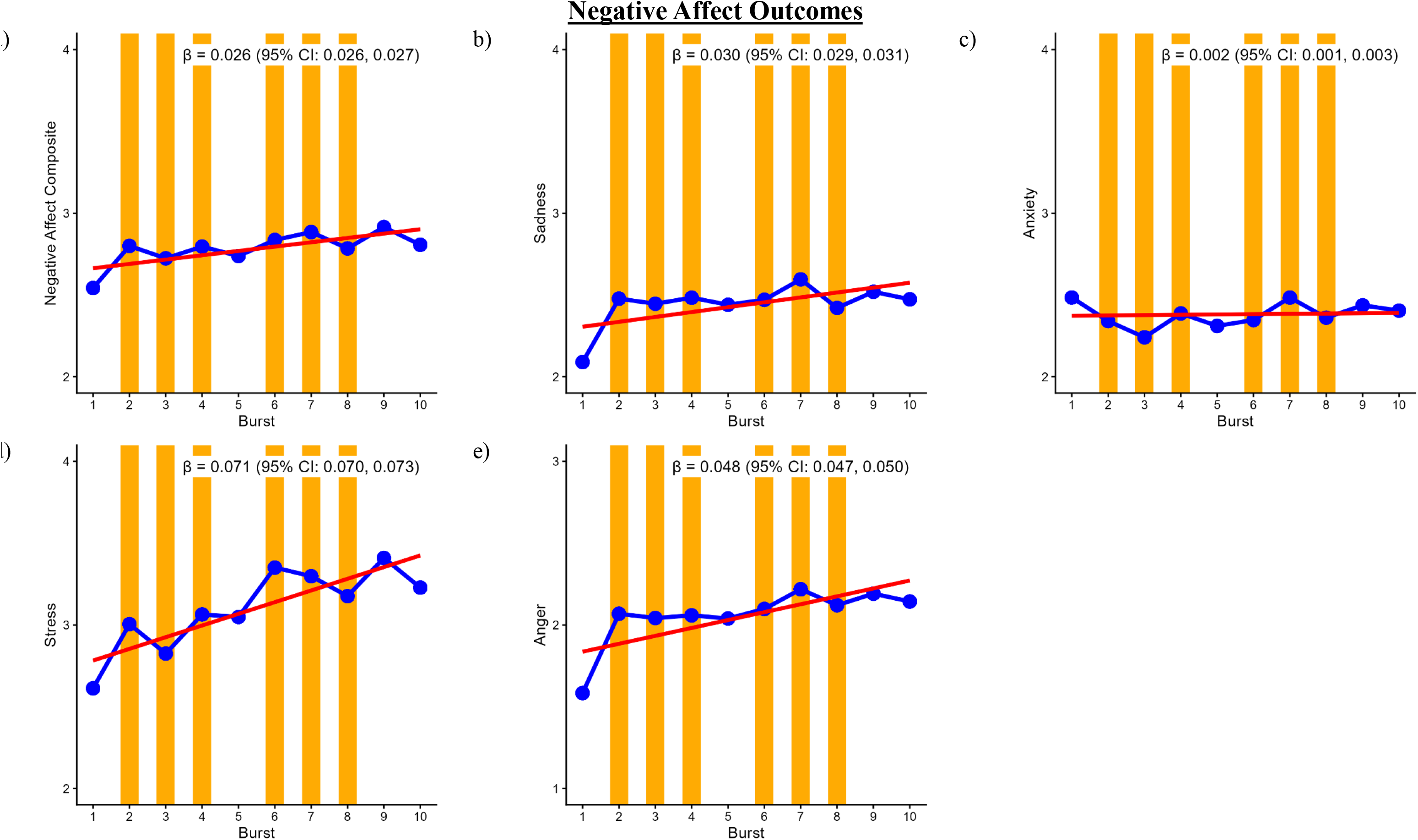
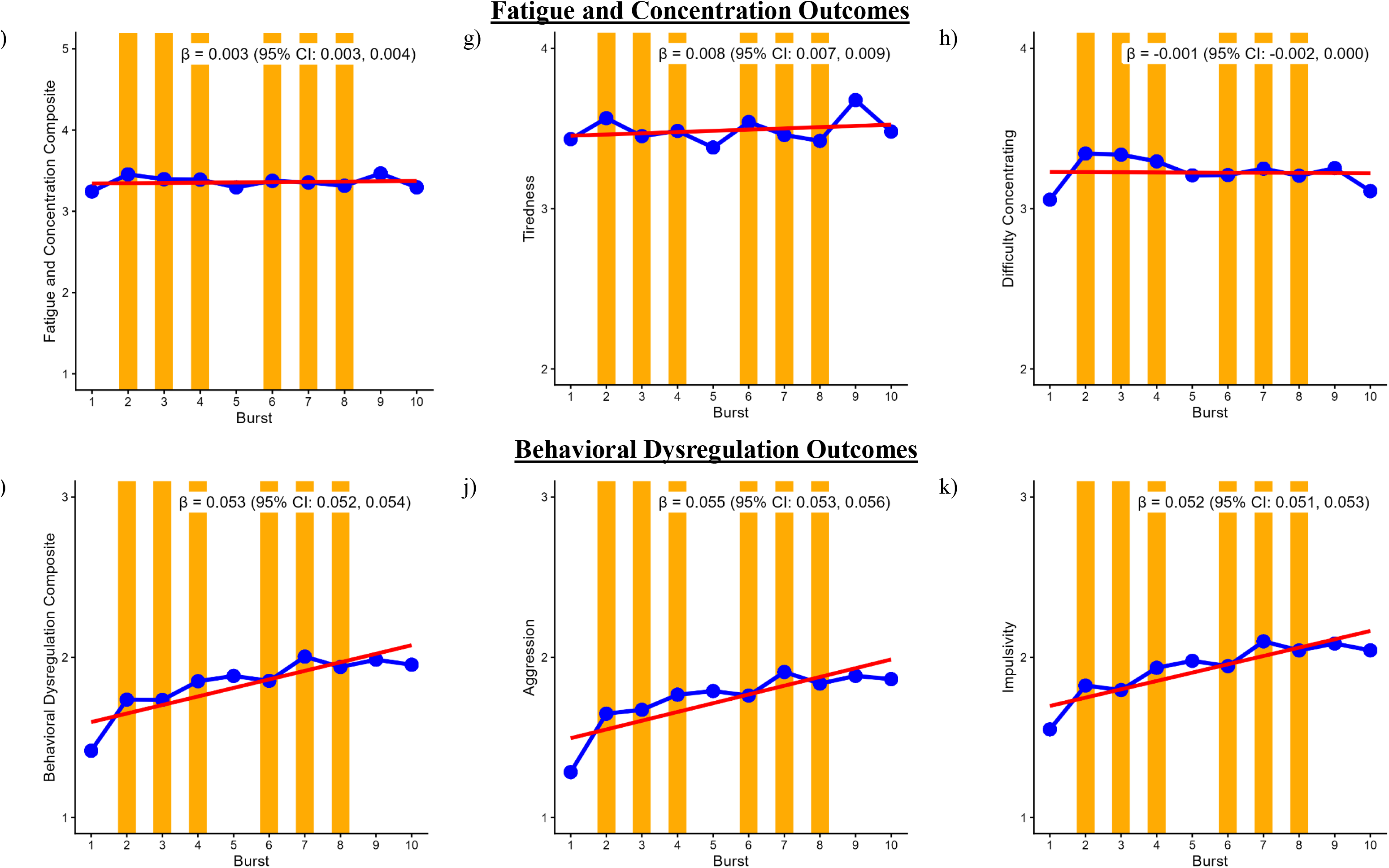
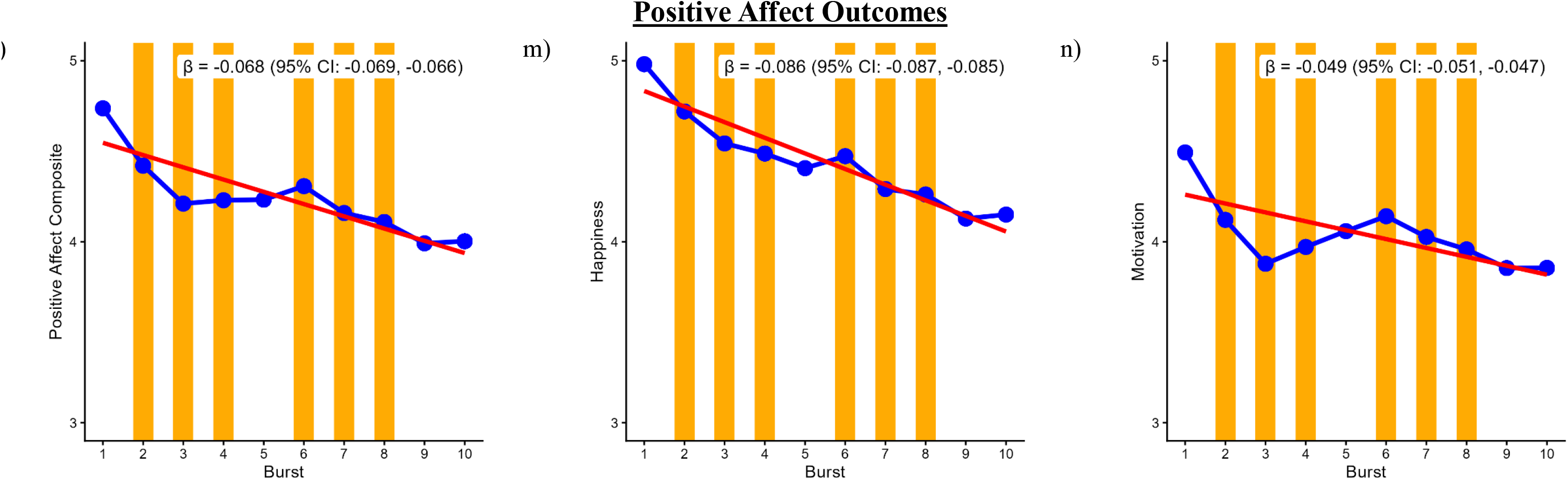
Mean mood scores by bursts and longitudinal association between burst number and mood for ESPINA-EMCT (n=496, nobs=14,332) from July 2022 to September 2024. Blue points and connecting lines represent the average sadness score at each EMA burst. The red regression line represents the estimated longitudinal association between burst number and sadness scores across the study period. Orange shaded regions indicate EMA bursts that occurred during peak pesticide spray season. Estimates are for the association between burst and averaged mood outcomes for each burst, adjusted for age, sex, race, monthly household income, participant education, and a participant random intercept.

GLMMs assessed the pooled and disaggregated (within-person and between-person) associations between peak versus non-peak pesticide spray season with mood outcomes, including domain-level (e.g., positive affect) and individual symptom (e.g. happiness) scores. Models adjusted for the same fixed and random effects as above, with the addition of a fixed effect assessment day within each burst (days 1-10) and a random intercept for burst nested within participant. For the pooled estimates, the independent variable was peak pesticide spray season, with non-peak seasons as the reference category.

To evaluate both the immediate and cumulative effects of pesticide spray season on daily mood outcomes, we disaggregated the categorical seasonal exposure variable (peak vs. non-peak) into independent *between-person* and *within-person* components. Because the study utilized a 3-level measurement burst design (daily observations nested within bursts, nested within participants) and seasonal exposure varied only at the burst level (bursts were either within peak or non-peak periods), the disaggregation was strictly calculated at the burst level. Calculating person-means using raw daily observations would artificially conflate a participant’s true environmental exposure with their daily EMA compliance rate (e.g., answering more surveys during a Peak burst would falsely skew their baseline exposure metric). To prevent this compliance bias, variables were derived by treating each burst as a single, equally weighted unit:

- *Between-Person Component:* Calculated as the proportion of total bursts each participant completed during the Peak season, which was subsequently grand-mean centered. This term evaluates whether individuals with a higher cumulative seasonal exposure burden across the study exhibited poorer overall mood levels compared to those with lower exposure.
- *Within-Person Component:* Calculated by subtracting the participant’s overall person-mean from the seasonal status of their current burst (1 = Peak, 0 = Non-Peak). This term isolates intra-individual effects and evaluates whether a participant experiences a significant shift in daily mood during a Peak burst relative to their own average mood during a Low burst.

Both the within-person and between-person components were included simultaneously as fixed effects in the GLMM, as well as nested random intercepts to account for the dependency of days within bursts and bursts within participants.

Effect modification by sex, floricultural occupation, and cohabitation with a floricultural worker was assessed by introducing an interaction term between the primary pesticide spray season variable and each respective covariate into the pooled GLMM. Statistically significant interaction terms were then stratified by the effect modifier.

All analyses were performed using R, version 2025.09.2. ChatGPT from Open AI (https://chatgpt.com/) was used to troubleshoot R coding errors during the data analysis process. All coding suggestions were reviewed and edited before implementation.

## Results

### Participant Characteristics

Of the 505 ESPINA-EMCT participants (Number of observations [Nobs]=16,154), 9 individuals and 1,822 observations were excluded due to missing EMA, missing covariates, or failing our alcohol and cannabis inclusion criteria (see Statistical Analysis section; Figure 1). Thus, a total of 496 unique participants and 14,332 EMA observations were included, totaling 5,113 non-peak and 9,219 peak spray season observations (Table 1). Mean age was slightly older during low spray seasons than during peak spray seasons (21.67 vs. 21.20 years, p<0.001). Peak spray season observations, versus non-peak observations, had a greater proportion of females (62.7% vs. 60.6%, p = 0.01) and floricultural workers (35.3% vs. 18.2%, p = 0.03), but a lower proportion of participants living with a floricultural worker (62.6% vs. 65.2%, p = 0.002). The first assessment, referred to as “Burst 1” for ease of explanation although it consisted of a single in-person administration, had a 100% completion rate. Average completion of remaining bursts, which were self-administered in participants daily lives, ranged from 53.7% for Burst 2 to 83.6% for Burst 10 (Table 2).

**Table 1.** Participant Demographics of unique participants, and total observations for the ESPINA-EMCT cohort.

|  | Unique<br>Participants | Total<br>Observations | Non-Peak<br>Spray<br>Season | Peak Spray<br>Season | p-<br>diff* |
| --- | --- | --- | --- | --- | --- |
| N | 496 | 14332 | 5113 | 9219 |  |
| Age (years) | 20.32 (1.81) | 21.37 (1.92) | 21.67 (1.94) | 21.20 (1.88) | <0.001 |
| Female (%) | 50.80% | 62.00% | 60.60% | 62.70% | 0.01 |
| Race |  |  |  |  | 0.12 |
| Indigenous | 18.50% | 14.10% | 14.70% | 13.80% |  |
| Mestizo, White or Other | 81.50% | 85.90% | 85.30% | 86.20% |  |
| Monthly Household Income (%) |  |  |  |  |  |
| Less than \$500 | 51.40% | 52.80% | 52.40% | 53.00% | 0.50 |
| \$500-\$999 | 22.80% | 23.60% | 24.10% | 23.30% | |
| \$1000+ | 25.80% | 23.60% | 23.40% | 23.70% | |
| Participant Education (years) | 12.56 (1.49) | 12.71 (1.47) | 12.72 (1.46) | 12.70 (1.48) | 0.52 |
| Works in Floriculture (%) | 18.80% | 19.40% | 18.20% | 35.30% | 0.03 |
| Lives with a Floricultural Worker (%) | 60.10% | 63.50% | 65.20% | 62.60% | 0.002 |

Values are percent (%) for categorical variables and mean (SD) for continuous variables.
\*p-diff was calculated using chi-square for categorical data, and ANOVA for continuous variables.

**Table 2.** Mean Adherence rates of EMA completion for each burst in the ESPINA-EMCT cohort (n=496, nobs=14,332) from July 2022 to September 2024.

| **Burst** | **Max EMAs** | **Mean Completed** | **Mean Adherence** | **N<sub>participants</sub>** |
| --- | --- | --- | --- | --- |
| 1 | 1 | 1 | 100% | 481 |
| 2 | 10 | 5.3 | 53% | 312 |
| 3 | 10 | 5 | 50% | 299 |
| 4 | 10 | 6.1 | 61% | 357 |
| 5 | 10 | 5.4 | 54% | 305 |
| 6 | 5 | 3.6 | 72% | 331 |
| 7 | 5 | 3.3 | 66% | 327 |
| 8 | 5 | 4 | 80% | 346 |
| 9 | 5 | 3.8 | 76% | 341 |
| 10 | 5 | 4.2 | 84% | 335 |
| Overall | 66 | 29 | 44% | 496 |

EMA=Ecological Momentary Assessment

### Mood Characteristics

Table 3 presents descriptive statistics for mood, aggregated at the unique participant level (N = 496) and the total observation level (n = 14,332). Overall, participants reported predominantly high levels of positive affect across the study duration. At the between-person level, positive affect had the highest overall average (Mean = 4.35, SD = 1.19), driven primarily by strong reports of feeling happy (Mean = 4.52, SD = 1.18). Fatigue and concentration followed with a mean of 3.25 (SD=1.34). In contrast, overarching participant-level averages for negative affect (Mean = 2.20, SD = 1.13) and behavioral dysregulation (Mean = 1.83, SD = 0.96) were lower, skewing toward the “not at all” end of the scale.

**Table 3.** Summary statistics for mood variables.

|  | Unique Participants<br>(N=496) |  | Total Observations<br>(N=14,332) |  |
| --- | --- | --- | --- | --- |
|  | Mean (SD) | Range | Mean (SD) | Range |
| Negative Affect | 2.20 (1.13) | 1 - 6.25 | 2.51 (1.38) | 1 - 7 |
| I feel stressed. | 3.07 (1.30) | 1 - 7 | 3.11 (1.86) | 1 - 7 |
| I feel angry. | 2.06 (1.03) | 1 - 6.48 | 2.08 (1.47) | 1 - 7 |
| I feel anxious. | 2.44 (1.17) | 1 - 6.61 | 2.37 (1.60) | 1 - 7 |
| I feel worried. | 2.82 (1.46) | 1 - 7 | 2.82 (1.79) | 1 - 7 |
| Behavioral Dysregulation | 1.83 (0.96) | 1 - 6.58 | 1.86 (1.29) | 1 - 7 |
| I feel aggressive. | 1.73 (0.94) | 1 - 6.58 | 1.77 (1.30) | 1 - 7 |
| I feel Impulsive. | 1.93 (1.06) | 1 - 6.58 | 1.95 (1.44) | 1 - 7 |
| Positive Affect | 4.35 (1.19) | 1.47 - 7 | 4.21 (1.60) | 1 - 7 |
| I feel happy. | 4.52 (1.18) | 1.80 - 7 | 4.42 (1.67) | 1 - 7 |
| I feel motivated. | 4.18 (1.35) | 1 - 7 | 4.00 (1.89) | 1 - 7 |
| Fatigue and Concentration | 3.25 (1.34) | 1 - 7 | 3.37 (1.55) | 1 - 7 |
| I am having trouble concentrating. | 3.15 (1.36) | 1 - 7 | 3.25 (1.80) | 1 - 7 |
| I feel tired. | 3.43 (1.68) | 1 - 7 | 3.49 (1.83) | 1 - 7 |

SD = Standard Deviation. Unique participants mean scores were calculated by first averaging each participant's responses for a given mood item across all their observations. These participant-specific means were then averaged across participants to obtain the overall mean score for unique participants. Summary statistics for total observations were calculated using all observation-level responses, with means derived by averaging each mood item across all observations.

Notably, the data showed systemic differences between the participant-level and the observation-level means, indicating differential response rates. For positive affect, the observation-level mean (Mean = 4.21, SD = 1.60) was noticeably lower than the participant-level mean. While the behavioral dysregulation (Mean = 1.86, SD = 1.29) and negative affect items (Mean = 2.79, SD = 1.32), were similar.

Across the bursts (Figure 2), there were increasing self-reports of all negative affect outcomes (Fig 2a-2e), tiredness (Fig 2g), and behavioral dysregulation outcomes (Fig 2i-2k). Conversely, scores of the positive affect composite (Fig 2l-2n) decreased over time.

### Peak Pesticide Spray Season and Mood Outcomes

#### Negative Affect (Figure 3)

Peak pesticide spray season was significantly associated with an overall increase in the negative affect composite (β = 0.13; 95% CI: 0.07, 0.18). When evaluating the individual mood variables within this domain, the strongest positive associations were observed for anger (β = 0.16; 95% CI: 0.10, 0.22), stress (β = 0.15; 95% CI: 0.07, 0.22), and sadness (β = 0.13; 95% CI: 0.07, 0.20). Associations did not reach statistical significance for anxiety.

**Figure 3.**
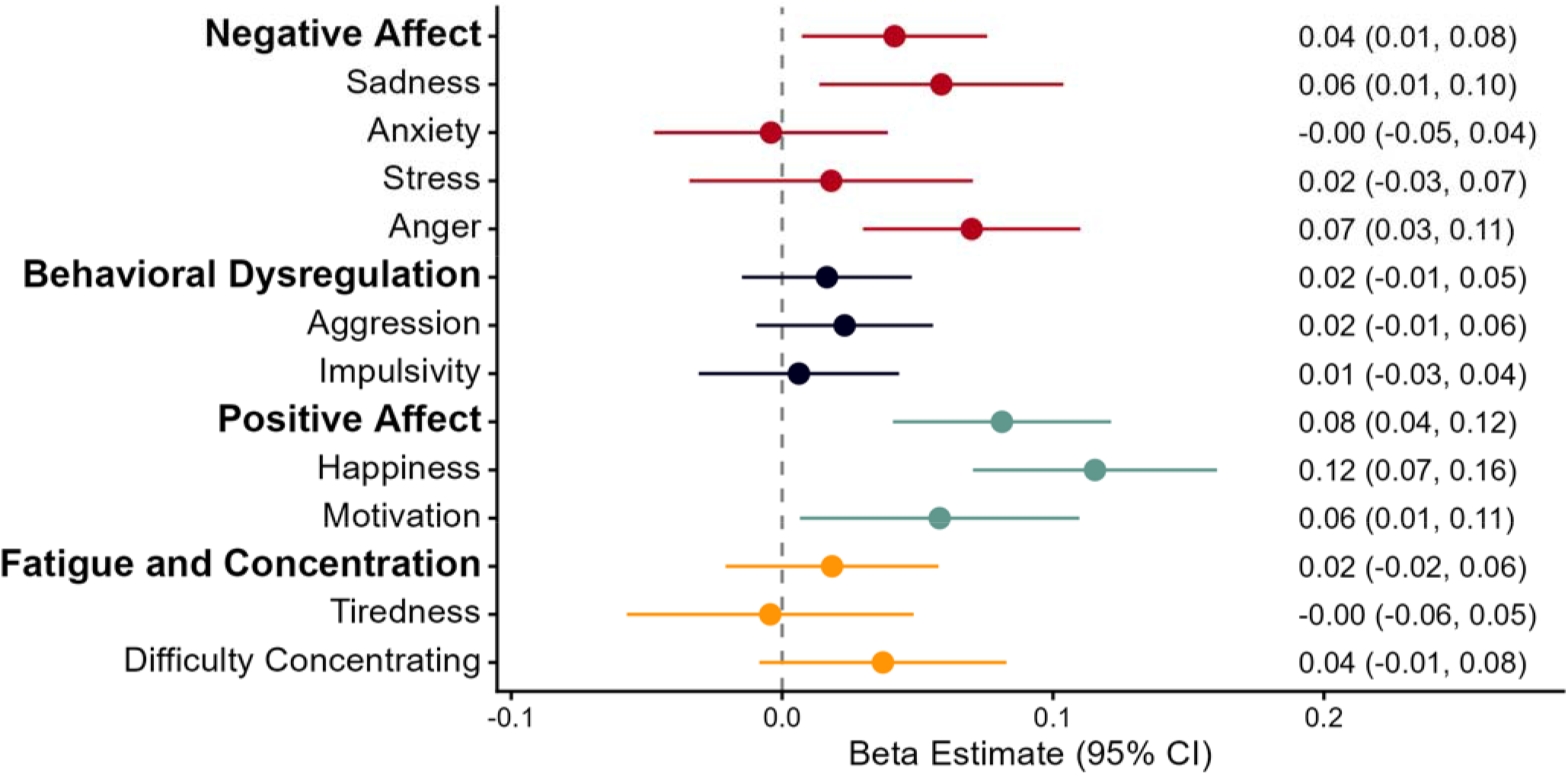
Associations between peak pesticide spray season and ecological momentary assessment (EMA)-derived mood outcomes among participants in the ESPINA-EMCT cohort (n=496, nobs=14,332) from July 2022 to September 2024. Models adjusted for assessment day within each burst (days 1–10), longitudinal burst number (bursts 1– 10), age, sex, race, monthly household income, participant education, participant random intercept, and participant nested burst random intercept. Mood domain scores were calculated by averaging component mood scores. Negative affect averaged reported feelings of sadness, anxiety, stress, and anger. Behavioral Dysregulation averaged reported feelings of aggression and impulsivity. Positive affect averaged reported feelings of happiness and motivation. Fatigue and Concentration averaged reported feelings of tiredness and difficulty concentrating

#### Behavioral Dysregulation (Figure 3)

A statistically significant positive association was observed between peak pesticide spray season and the overall behavioral dysregulation composite (β = 0.10; 95% CI: 0.05, 0.15). Both individual variables of this domain demonstrated similar significant increases in peak pesticide spray seasons (aggression (β = 0.10; 95% CI: 0.05, 0.15) and impulsivity (β = 0.09; 95% CI: 0.04, 0.15)).

#### Positive Affect (Figure 3)

In contrast to the negative mood domains, peak pesticide spray season was not significantly associated with the positive affect composite (β = 0.01; 95% CI: −0.05, 0.07). The individual variables (happiness, motivation) similarly demonstrated null associations with the peak spray season.

#### Fatigue and Concentration (Figure 3)

Peak pesticide spray season was associated with greater overall fatigue and concentration difficulty on the composite measure (β = 0.06; 95% CI: 0.001, 0.12), but not significantly associated with tiredness or difficulty concentrating when examined separately.

#### Disaggregated Within and Between Person Associations

Associations between peak pesticide spray season and EMA-derived mood outcomes varied between the within-person and between-person estimates (Figure 4). *Within-person associations* indicated that participants reported worse mood during their own peak spray season periods compared with their own low spray season periods. Peak spray season was associated with higher negative affect (β = 0.13 [0.07, 0.18]), driven by higher sadness (β = 0.13 [0.07, 0.19]), stress (β = 0.15 [0.07, 0.22]), and anger (β = 0.16 [0.10, 0.22]). It was also associated with greater behavioral dysregulation (β = 0.10 [0.05, 0.15]), including both aggression (β = 0.10 [0.05, 0.15]), and impulsivity (β = 0.09 [0.03, 0.15]) scores. Positive affect as well as fatigue and concentration variables were not associated with peak pesticide season.

**Figure 4.**
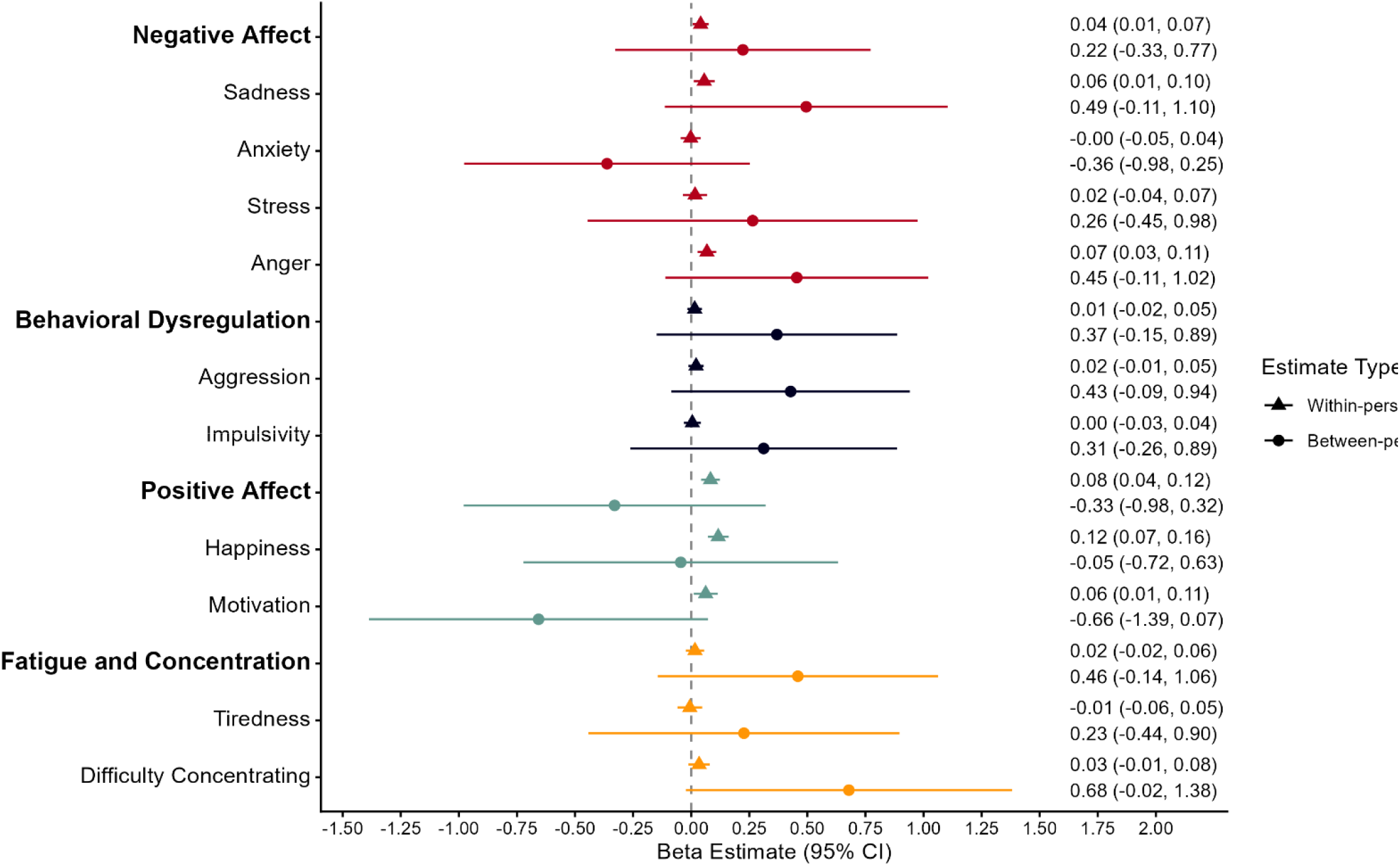
Within-person and between-person associations between peak pesticide spray season and ecological momentary assessment (EMA)-derived mood outcomes among participants in the ESPINA-EMCT cohort (n=496, nobs=14,332) from July 2022 to September 2024. Models adjusted for assessment day within each burst (days 1–10), longitudinal burst number (bursts 1– 10), age, sex, race, monthly household income, participant education, participant random intercept, and participant nested burst random intercept. Mood domain scores were calculated by averaging component mood scores. Negative affect averaged reported feelings of sadness, anxiety, stress, anger, tiredness, and difficulty concentrating. Behavioral Dysregulation averaged reported feelings of aggression and impulsivity. Positive affect averaged reported feelings of happiness and motivation. Fatigue and Concentration averaged reported feelings of tiredness and difficulty concentrating

In contrast, *between-person associations* were substantially less precise for all outcomes. Although point estimates for negative affect-related outcomes and behavioral dysregulation were positive, all confidence intervals included the null, except for difficulty concentrating, which demonstrated the largest between-person estimate (β = 0.71 [0.001, 1.41]). Peak spray season remained non-significant with positive affect, as well as fatigue and difficulty concentrating.

### Effect modification by Floricultural Occupational Status and Cohabitation with a Floricultural Worker

We observed effect modification between peak pesticide spray seasons with floriculture occupational status for positive affect (β_interaction_=-0.25 [−0.42, −0.06], p=0.004), and stratified analyses by floricultural occupation status are presented in Figure 5. Among floricultural workers, there were negative associations between peak pesticide spray season and positive affect (β = −0.16, [−0.32, 0.00]), but not in non-occupational participants. Mood and behavioral dysregulation outcomes were generally poorer among floricultural workers compared with non-floricultural workers. At the component level, peak pesticide spray seasons were associated with greater sadness (β = 0.21, 95% CI: 0.02, 0.40) and stress (β = 0.25, 95% CI: 0.04, 0.47) in floricultural versus non-floricultural workers. For the behavioral dysregulation domain, peak spray seasons were associated with higher composite behavioral dysregulation (β = 0.14, [−0.01, 0.28]), driven by higher impulsivity scores (β = 0.16, [0.00, 0.32]) among floricultural workers. Effect modification by cohabitation with a floricultural worker was not observed.

**Figure 5.**
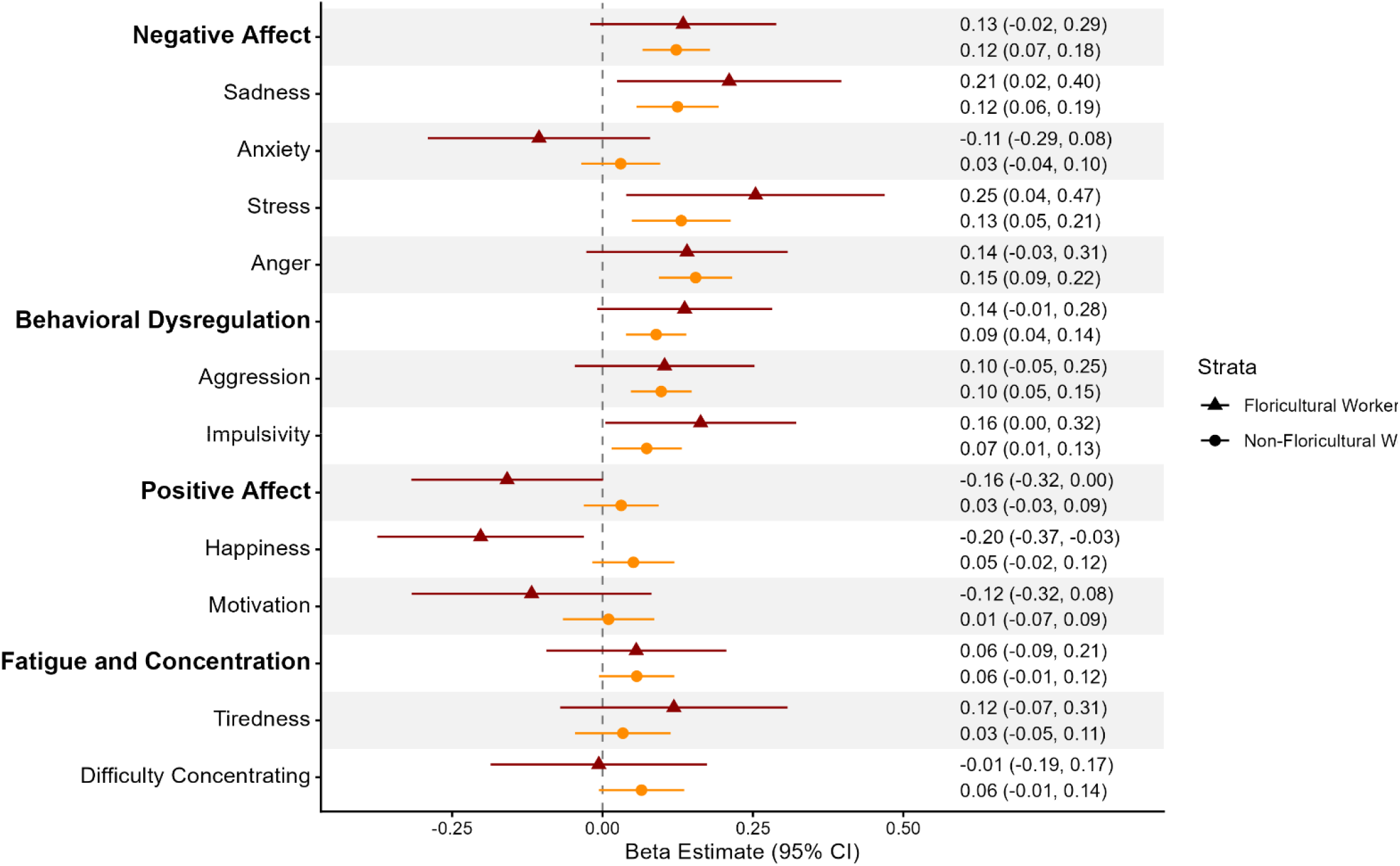
General Linear Mixed Effect Associations between peak pesticide spray season with mood, stratified by floricultural occupation status in the ESPINA-EMCT cohort (n=496, nobs=14,332) from July 2022 to September 2024. Models adjusted for assessment day within each burst (days 1–10), longitudinal burst number (bursts 1– 10), age, sex, race, monthly household income, participant education, participant random intercept, and participant nested burst random intercept.

#### Effect modification by Sex

There was limited evidence of effect modification by sex. A significant interaction between peak pesticide spray season and sex was observed for anger (β_interaction_ =0.12 [0.01, 0.23]; p=0.04). In sex-stratified analyses, associations during peak pesticide spray seasons were stronger among males than females for anger (β_boys_=0.24 [0.16, 0.33]vs β_females_=0.11 [0.03, 0.19]).

## Discussion

This study found peak pesticide spray seasons were consistently associated with greater negative affect and behavioral dysregulation among adolescents and young adults living in an intensive floricultural region of Ecuador. Using repeated ecological momentary assessments collected over a two-year period, we observed primarily within-person associations between peak spray seasons and higher reported sadness, stress, anger, aggression, and impulsivity. These findings suggest that seasonal increases in pesticide use may contribute to short-term fluctuations in mood. Importantly, the predominance of within-person associations indicates these mood changes occurred within the same individuals across spray seasons, strengthening evidence that pesticide exposure periods may influence day-to-day emotional well-being rather than simply reflecting stable differences between participants.

Our findings are consistent with epidemiologic literature linking pesticide exposure with depression, anxiety, and broader neurobehavioral dysfunction. Previous studies among agricultural workers have demonstrated greater odds of depression following pesticide poisoning events ^9^, and systematic reviews have found that the majority of studies report positive associations between pesticide exposure and depressive symptoms^10^. Similar findings have also been observed in para-occupational populations, including among spouses of pesticide applicators ^11^ and children living with floricultural workers ^13^. Our findings extend this literature by demonstrating that mood symptoms may fluctuate alongside seasonal pesticide application patterns in real-world settings, even outside of acute poisoning events. The contrast between the within-person and between-person findings suggests that pesticide-related mood effects may be driven primarily by temporal changes in exposure rather than by stable differences between individuals. While participants generally experienced greater negative affect and behavioral dysregulation during peak spray seasons, individuals with higher average exposure to spray periods across the study period did not consistently exhibit worse mood than those with lower average exposure. This finding suggests that seasonal increases in pesticide exposure may trigger short-term neurobehavioral responses that fluctuate over time, rather than reflecting solely chronic differences between participants.

The strongest and most consistent associations were observed for negative affect, particularly sadness, stress, and anger. In contrast, positive affect was not associated with peak spray seasons overall. Peak spray season was also associated with slightly greater overall fatigue and concentration difficulty on the composite measure, although associations with tiredness and difficulty concentrating were not significant when examined separately. This suggests that periods of elevated pesticide exposure may have subtle effects on perceived cognitive or physical functioning in addition to mood and behavioral regulation. The contrasting findings for negative and positive affect may indicate that pesticide-related neurobehavioral effects manifest more strongly through increases in negative emotional states rather than reductions in positive affect. Negative affect may also be more sensitive to short-term environmental stressors and neurotoxic exposures than broader measures of positive mood. Differential impacts in negative and positive affect have been reported for characteristics like age, where individuals of older age have lower mean levels and shorter duration of negative emotional experiences^52^. Negative and positive affect have also been found to be independent of each other^6^.Together, these findings suggest distinct biological and psychological mechanisms may underlie positive and negative affective responses to environmental exposures. Across time, we also observed greater negative affect and behavioral dysregulation, but lower positive affect scores. These patterns may reflect the cumulative effects of chronic pesticide exposure resulting from repeated seasonal exposures over many years. While peak spray seasons likely capture periods of heightened exposure, participants living in this agricultural region may also experience ongoing low-level exposures throughout the year. Such repeated exposures could contribute to persistent disruptions in neurobiological processes involved in mood regulation, emotional processing, and executive functioning.

Behavioral dysregulation outcomes, consisting of aggression and impulsivity, were elevated during peak spray seasons. While much of the pesticide literature focuses on depression and anxiety, fewer studies have evaluated emotional regulation and impulsive behaviors directly. Evidence from animal models is consistent with our findings. For impulsivity, rats that were administered a dose of 250 mg/kg of chlorpyrifos demonstrated more impulsive choices at 10 weeks and 1 year after administration ^53^. For aggression, repeat exposure to methamidophos, an organophophorus pesticide, induced attached behavior in non-aggressive mice, suggesting it may have a pro-aggressive effect ^54^. Exposing native (*Monomorium antarcticum*) and invasive ant species (*Linepithema humile*) to neonicotinoids influenced aggressive dynamics between the species^55^. In humans, one potential mechanism is impairment of inhibitory control, a core executive function that enables individuals to suppress inappropriate thoughts, actions, or responses and regulate behavior ^56^. Past ESPINA studies have consistently demonstrated that higher pesticide exposure, as indicated by higher urinary pesticide metabolite concentrations, lower AChE activity, and less time after a the Mother’s Day harvest, was associated with poorer inhibitory control ^29,36,57,58^. These findings suggest that pesticide exposures may contribute to increased impulsive behavior through disruption of neurocognitive processes involved in self-regulation and behavioral control.

Although positive affect was not associated with spray season overall, floricultural workers experienced lower positive affect during peak spray seasons compared with non-occupational participants. These findings may reflect greater direct occupational exposure among workers during periods of intense agricultural production. This finding is consistent with other studies compared showing that agricultural workers have had higher exposure to pesticides indicated by urinary metabolite concentrations ^33,59–62^, AChE activity ^59^, and butrylcholinesterase activity ^59^. Likewise, floriculture extends beyond economic activity, but also shapes families, work and community dynamics, which may influence lower positive affect amongst workers. However, we did not observe effect modification by cohabitation with a floricultural worker. This may be indicative that additional exposure from para-occupational pathways may not be sufficient to affect mood alterations.

There was limited evidence of effect modification by sex, where associations between peak spray seasons with anger were stronger among males compared to females, but not for other mood outcomes. In ESPINA, we have previously reported stronger depression symptom associated with pesticide exposure in adolescent females ^13^ but stronger associations among adolescent males for the neurobehavioral domains of attention, inhibitory control, and visuospatial processing ^29^. This highlights the possibility that pesticide-related exposures may preferentially affect executive functioning and behavioral regulation among males, contributing to greater anger during periods of increased agricultural activity. Endocrine alterations may underly our findings. We have found that greater insecticide exposure was positively associated with testosterone, 17β-Estradiol, and DHEA concentrations, with stronger associations in male adolescents ^63^. In the same time period in boys, elevated estradiol and testosterone was positively associated with elevated depression scores ^26^. More research is needed to better understand the underlying mechanism.

This study has several important strengths. To our knowledge, this is the largest study to date to assess the relationship between pesticide exposure periods and daily mood fluctuations. Leveraging a longitudinal burst design, we captured an exceptionally robust 14,332 observations from a substantial sample of 496 participants, providing the statistical power necessary to disaggregate the variance and reliably detect subtle, short-term, within-person fluctuations in mood that would be masked in smaller or traditional cross-sectional designs. Second, using repeated, real-time EMA measurement of mood in participants’ natural environments, reduces recall bias and improves ecological validity compared to traditional retrospective questionnaires. The longitudinal burst design further enabled assessment of both between-person and within-person changes across multiple spray seasons over two years. Finally, conducting this study within a well-characterized longitudinal cohort in a region with highly predictable, cyclical pesticide spray cycles provided a unique opportunity to reliably isolate the temporal relationships between agricultural production peaks and emotional well-being. Tracking the same individuals across alternating periods of high and low agricultural production provides a much stronger temporal framework for evaluating acute/subacute effects than single-timepoint measurements.

Several limitations should be considered. First, pesticide exposure was inferred using peak pesticide spray seasons rather than biomarker measurements collected concurrently with each EMA assessment. Although prior studies in this population have demonstrated higher pesticide exposure biomarkers during peak floricultural production period ^29,34,35^, exposure misclassification remains possible because this proxy does not capture individual variability in occupational activities, residential proximity, or protective behaviors. Such non-differential misclassification would likely bias estimates toward the null. Second, mood was self-reported and may be influenced by unmeasured psychosocial or contextual factors and are not formal diagnostic evaluations. We are unable to comment on the finding’s implications on mental health conditions, however, under the tripartite model negative affect has been connected with both depression and anxiety. Although the EMA minimizes recall bias by capturing real-time, repeated appraisals, it remains vulnerable to subjective interpretation. Furthermore, intensive repeated sampling can introduce participant survey fatigue or reactivity, although this is a common trade-off in high-frequency longitudinal designs. Third, residual confounding by factors such as stressors, sleep, workload, or economic conditions may persist^64,65^. Finally, these findings may not generalize beyond agricultural communities engaged in intensive floricultural production. Floriculture is predominantly conducted within enclosed greenhouse environments, thus community pesticide drift may be lower than in regions dominated by open-field agriculture or aerial spraying, where associations between pesticide exposure and mood outcomes could be even stronger.

## Conclusion

Peak pesticide spray seasons were associated with transient increases in negative affect and behavioral dysregulation among adolescents and young adults living in an agricultural community. We also observed a small increase in a composite measure of fatigue and concentration difficulty, suggesting that these short-term effects may extend beyond emotional functioning. These findings suggest that seasonal intensification of pesticide use may influence day-to-day emotional functioning and highlight the importance of considering temporal variability in pesticide exposure when evaluating mental health outcomes. EMA approaches may provide a sensitive framework for identifying transient neurobehavioral impacts of environmental exposures and could help inform interventions aimed at reducing mental health risks during periods of elevated pesticide use. These results highlight the need to implement prevention, exposure monitoring, and health protection strategies tailored to the cultural and community context of floricultural regions.

## Data Availability

All data produced in the present study are available upon reasonable request to the authors

## Acknowledgments

We extend our sincere gratitude to the ESPINA study staff, Fundación Cimas del Ecuador, the Parish Governments of Pedro Moncayo County, the community members of Pedro Moncayo, and the Education District of Pichincha-Cayambe-Pedro Moncayo for their invaluable support and contributions to this project. We would like to thank Rajendra Parajuli for his help in the preparation of submission materials.

